# A Multistage Mixed Methods Methodology to Develop a Toolkit to Reduce Infections Following Durable LVAD Implantation

**DOI:** 10.1101/2025.02.24.25322824

**Authors:** P. Paul Chandanabhumma, Sriram Swaminathan, Lourdes Cabrera, Shiwei Zhou, Carol E. Chenoweth, Hechuan Hou, Sarah Comstock, Preeti N. Malani, Keith D. Aaronson, Francis D. Pagani, Donald S. Likosky, the Michigan Congestive Heart Failure Investigators

## Abstract

**Background:** Infections following durable left ventricular assist device (dLVAD) implantation are common and associated with increased morbidity and mortality. Despite documented interhospital variability, few studies have identified strategies to mitigate their occurrence. This national study uses a multistage mixed methods design to develop a customizable and deployable toolkit of expert-guided recommendations to reduce infections post-dLVAD.

**Methods:** Representatives from low, medium, and high-performance hospitals across the U.S. were interviewed to assess factors contributing to post-dLVAD infections. Draft toolkit recommendations were iteratively developed after integrating thematically analyzed qualitative and quantitative data. A national advisory team of ventricular assist device (VAD) subject matter experts provided mixed methods input to refine the toolkit’s content and structure.

**Results:** Seventy-three clinical and operational VAD team members were interviewed, and 14 subject matter experts provided stakeholder feedback to refine the toolkit. The resulting toolkit contains 39 infection prevention recommendations that address VAD program care processes (e.g., real-time provider communication), clinicians (e.g., multidisciplinary protocol development), patients & caregivers (e.g., engaging patient advisors in patient education), and VAD leadership (e.g., unit and service level data reporting). Accompanying resources (e.g., team-based exercises, data collection worksheets) support implementing and evaluating site-specific strategies. Input from clinical and research experts provided preliminary evidence of the toolkit’s acceptability and considerations for enhancing the toolkit’s adoption and implementation.

**Conclusions:** Using mixed methods approaches, an infection prevention toolkit was developed to enhance care coordination among VAD team members and mitigate post-dLVAD infections. Future work should evaluate the effectiveness of implementing this infection prevention toolkit within the dLVAD setting.

**What is Known:**

- Six out of every 10 patients develop an infection within 2 years of durable left ventricular assist device (dLVAD) implantation.
- Infections contribute to an increased risk of major morbidity and mortality.
- Post-dLVAD infection rates vary across hospitals, even after risk adjustment.

**What the Study Adds:**

- A customizable toolkit of infection prevention recommendations was developed and pilot tested using mixed methods approaches.
- Recommendations focus on modifiable aspects of organizational structure, patient care, patient education, quality reporting.

## INTRODUCTION

Patients undergoing durable left ventricular assist device (dLVAD) are at risk for developing post-implantation infections.^1^ Analyses of the Multicenter Study of MagLev Technology in Patients Undergoing Mechanical Circulatory Support Therapy with HeartMate 3 (MOMENTUM 3) clinical trial found that nearly 6 out of every 10 patients undergoing implant of a dLVAD develop a device-related infection (i.e., percutaneous lead) and/or non-device-related infection (i.e., pneumonia) within 2 years of implantation.^2^ These infections, which vary between 0.0 to 35.6 per 100 patient-months across hospitals,^3^ expose patients to an increased risk of major morbidity and resource utilization, diminishment in quality of life, and increased risk of mortality.^4–6^

Despite documented large-scale interhospital variability in infections, few studies have identified strategies to mitigate their occurrence.^7^ Early infection prevention efforts focused on identifying risk factors across phases of longitudinal care (e.g., pre-, intra- and post-operative settings) for patients with implanted dLVADs.^1,8,9^ Differences in infection rates across hospitals are minimally accounted for by patient risk.^3^ Therefore, efforts to advance infection prevention should evaluate other determinants, including the context (e.g., social determinants of health, caregiver support) and organizational structure (e.g., staffing and leadership model of the VAD program) of care, both within and outside of the index admission.

This multistage, mixed methods study was undertaken to develop a customizable and deployable toolkit of expert-guided infection prevention recommendations across the continuum of dLVAD care. The study team hypothesized that integrating VAD team members’ perspectives across hospitals and actively engaging key dLVAD subject matter experts would advance a customizable infection-prevention toolkit that may potentially reduce infections following dLVAD implantation.

## MATERIALS AND METHODS

### Overview

The study team grounded its approach towards creating the dLVAD Care Conceptual Framework based on: (1) the Joint Commission Patient Tracer Methodology;^10^ (2) prior literature findings;^1,8,9^ and (3) evolving analytical insights.^1,8,9^ The study’s framework (**eFigure 1**) delineates the phases of dLVAD care (e.g., pre-admission through post-discharge), key stakeholders involved (e.g., patients and caregivers, VAD coordinators, dLVAD surgeons, heart failure cardiologists, intra and postoperative nursing staff), and contextual domains of infection prevention (e.g., culture of collaboration, feedback reporting, data quality).

Methodologically, this study temporally consisted of three stages (**Figure 1)**.^7,11,12^ Stage 1 involved mixed methods formative assessment research. The assessment combined an explanatory sequential design with a case series approach. Specifically, the collection and analysis of quantitative data informed a holistic qualitative approach that focused on understanding care practices and experiences among hospitals that were each evaluated as complete cases (i.e., distinct organizational unit).^13^ The study team conducted (1) quantitative analyses of merged national datasets of dLVAD procedures to identify low and high-performing hospitals for a qualitative case series investigation and (2) qualitative in-depth interviews with clinical and administrative team members across the continuum of hospital performance to advance candidate infection prevention recommendation topics. During Stage 2, the study team integrated qualitative and quantitative findings and insights from Stage 1 to contextualize key determinants of hospital performance. Stage 3 involved developing, iteratively enhancing, and piloting the prototype toolkit based on clinical expert feedback from a national dLVAD Toolkit Advisory Team (dLVAD-AT) of multidisciplinary subject matter experts. Additionally, the toolkit was piloted for usability and intended effectiveness. All study phases received approval from the Institutional Review Board of Michigan Medicine (Study IDs: HUM00155687, HUM00165178, and HUM00218501).

**Figure 1.**
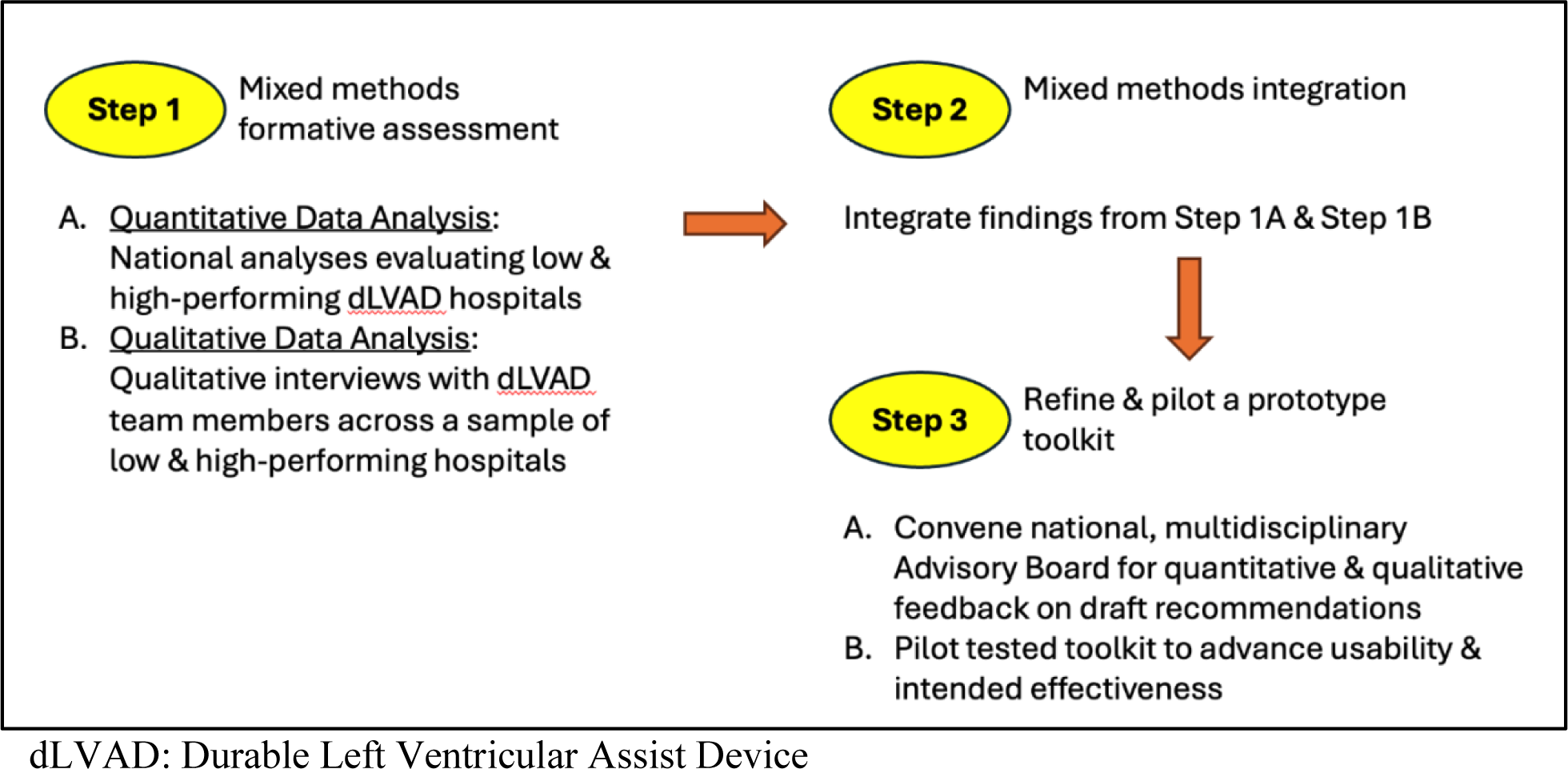
Study procedural diagram.

### Data Sources

The study leveraged several quantitative and qualitative data sources (**eTable 1)**. Low and high-performing case hospitals were identified for qualitative inquiry through the analysis of a merged national dataset comprising: (1) Interagency Registry for Mechanically Assisted Circulatory Support (Intermacs); (2) Center for Medicare & Medicaid Services (CMS) Fee-For-Service Medicare claims; and (3) American Hospital Association Annual Survey.^14^ Intermacs data were: (1) provided by the Intermacs Data Coordinating Center with permission from the National Heart, Lung, and Blood Institute before the administration of Intermacs was transferred to The Society of Thoracic Surgeons in January 2018 (previously funded, in part, by the National Heart, Lung, and Blood Institute, under Contract No. HHSN268201100025C). Registrant informed consent participation in the Intermacs data collection was required until Protocol v4.0 implementation (February 27, 2014).

Several data sources were used to advance stakeholder input to advance the toolkit development during Stage 1.^11^ Interviews were conducted with 73 participants across 8 U.S. hospitals that were selected for case series investigations based on their risk-adjusted, 180-day post-implantation infection rate. Four of the eight hospitals involved in the interviews were classified as high-performers (i.e., lowest tertile of risk-adjusted 180-day infection rate) while three hospitals were low-performers (i.e., highest tertile of infection rate) and one hospital was a middle-performer. Qualitative and quantitative feedback was received from the dLVAD-AT, while qualitative data was received during the pilot testing of the toolkit. Data use agreements restrict the distribution of raw study-related data files. Requests for summary statistics will be shared upon review and approval by the study team.

### Analysis and Toolkit Synthesis

During Stage 2, the analytical team (SS, PPC, LC, DL) iteratively developed the toolkit in collaboration with the broader study team. First, the Qualitative Analyst (SS) conducted thematic analyses of the semi-structured interviews, focusing on infection prevention roles, experiences, and recommendations.^15,16^ Analyses were also undertaken to identify thematic similarities and differences across quantitatively derived low- and high-performing hospitals and intended stakeholders (e.g., dLVAD surgeon, HF cardiologist)^17^. As noted above, the qualitative findings of all case hospitals were considered equally regardless of their infection performance.^13^ The drafted toolkit included the infection-prevention recommendations and topic-specific supporting materials that were inductively derived through qualitative and quantitative analyses. The drafted recommendations were stratified by care delivery domain, phase of dLVAD care, and intended stakeholder.

### Expert Feedback, Refinement, and Pilot Testing of the Toolkit

During Stage 3, the analytical team (SS, PPC, LC, DL) conducted several rounds of multi-method feedback to iteratively refine the toolkit recommendations, supporting materials, and implementation resources. The clinical and quantitative team members prioritized feedback, including but not limited to ranking the relative importance of each drafted recommendation and editorial suggestions to the toolkit statements. The analytical team developed criteria (see **eTable 2**) to consider editorial suggestions (e.g., syntax and clarification) for each recommendation, and more broadly discussed recommendation revisions with the study team.

Next, the analytical team conducted two rounds of focus group engagement with the dLVAD-AT. The dLVAD-AT consisted of a purposive sample of multidisciplinary subject matter experts. The dLVAD-AT members were affiliated with four hospitals (2 low-performing, 1 middle-performing, 1 high-performing). Subject matter experts agreed to provide quantitative (ratings of prototype recommendation content and phrasing) and qualitative (open-ended comments) survey feedback before each focus group. The 10-item surveys incorporated Likert responses: 1=Strongly agree; 2=agree; 3=neutral; 4=disagree, 5=strongly disagree. Using a “think out loud” approach, the focus groups involved soliciting feedback concerning the toolkit’s: (1) content, usability, acceptability, and likelihood of impact (during the first round of engagement); and (2) anticipated resource needs (e.g., personnel) as well as facilitators and barriers for implementing the proposed toolkit (during the second round of engagement). Using the revision approach described above, the quantitative and qualitative feedback was integrated before and after each round to enhance all toolkit components.

The toolkit was piloted at a middle-performance hospital. The pilot focused on gaining feedback on the toolkit’s usability and likelihood of success. The institution’s VAD Program Manager received feedback electronically and through discussions during dLVAD team meetings. The qualitative feedback was reviewed and incorporated into an iteratively enhanced toolkit.

## RESULTS

This mixed methods study involved 73 VAD team members from eight hospitals (interviews, Stage 1) and 14 dLVAD-AT participants (focus groups, Stage 2). The specialties and roles of the subjects are listed in **Table 1**.

**Table 1.** Characteristics of Participants.

|  | dLVAD Interview Participants<br>(n=73) |  |  | dLVAD<br>Toolkit<br>Advisory<br>Team (n=14) |
| --- | --- | --- | --- | --- |
|  | High<br>Performin<br>g<br>Hospitals | Middle<br>Performing<br>Hospitals | Low<br>Performing<br>Hospitals |  |
| Participant Center Volume,<br>median (IQR) | 33 (32-96) | 189 (189-189) | 192 (169-213) |  |
| 90-day Infection Rate per 100<br>patient-months | 2.52 | 5.93 | 26.18 |  |
| **Role** |  |  |  |  |
| <u>Physicians</u> |  |  |  |  |
| Cardiac Surgeon | 3 | 1 | 3 | 2 |
| Heart Failure Cardiologist | 3 | 1 | 5 | 2 |
| Hospital Epidemiologist | 0 | 1 | 3 | 1 |
| Anesthesiologist/Intensivist | 2 | 1 | 2 | 2 |
| Infectious Disease Specialist | 2 | 1 | 2 | 1 |
| <u>Advance Practice Provider</u> |  |  |  |  |
| Outpatient Advance Practice<br>Provider | 2 | 1 | 1 | 0 |
| Inpatient Advanced Practice<br>Provider | 2 | 1 | 3 | 0 |
| <u>RN</u> |  |  |  |  |
| Cardiac Surgery ICU Nurse | 1 | 1 | 2 | 1 |
| Ward Nurse | 2 | 0 | 3 | 0 |
| VAD Coordinator | 4 | 1 | 3 | 2 |
| Infection Prevention Specialist<br>Nurse | 0 | 2 | 2 | 2 |
| Quality Officer Nurse | 1 | 1 | 0 | 0 |
| Nurse Specialist | 2 | 0 | 1 | 0 |
| Floor Nurse | 0 | 0 | 0 | 1 |
| <u>Education Lead</u> | 0 | 0 | 0 | 2 |
| <u>LVAD Specialist</u> |  |  |  |  |
| Infection Prevention Specialists,<br>Perfusionists, etc. | 2 | 0 | 6 | 0 |

\*Durable Left Ventricular Assist Device (dLVAD)

Findings derived from analyses of the qualitative interviews supported the development of 19 infection prevention themes (**Table 2)**. Broadly, the themes intersected multiple levels and factors (e.g., patient, clinician, hospital) underlying infection risk, VAD infection prevention processes (e.g., patient selection, patient education, intra-operative care, post-discharge care, infection surveillance, provider and patient education), and facilitators and barriers to infection prevention adherence. Initially, 23 preliminary recommendations were drafted from these themes and subsequently categorized by context and anticipated stakeholder group. The contextual domains reflected the recommendation’s focus on <u>infection prevention</u> (e.g., culture of collaboration, feedback reporting, data quality) and phase of dLVAD care (e.g., pre-implantation to post-discharge, and longitudinal phase of care). Each recommendation had distinct goals, required resources and implementation requirements. As such, each recommendation was aligned with any of the four target stakeholder groups: VAD Program, Clinicians, Patients and caregivers, and VAD leadership.

**Table 2.**
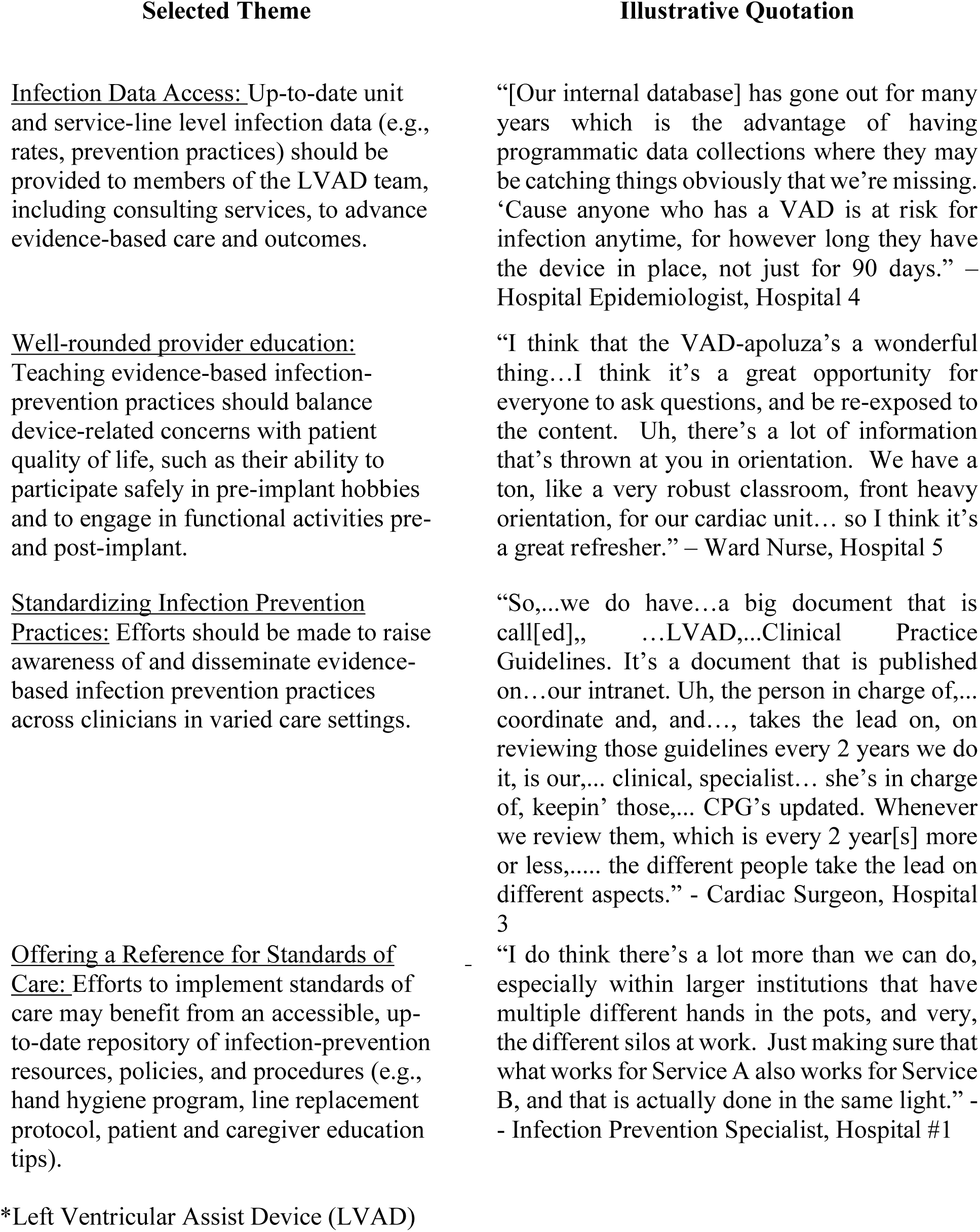
Illustrative Themes and Quotations from the Qualitative Interviews.

The recommendations were further edited by the substantive feedback from both team members and the dLVAD-AT focus groups. For instance, approximately a third of the recommendations received syntax and clarifying revisions. A fourth of the recommendations received contextual clarification, several recommendations were either split or combined, and two of the items were preserved in their initial form. Consequently, the total number of recommendations grew from 23 to 39. Illustrative examples of the iterative nature of the recommendations are documented in **eTable 2**.

The focus groups generated feedback related to the toolkit’s organization, supporting materials, and method of dissemination. Some of the input resulted in including: 1) an executive summary; 2) implementation materials to support team meeting discussions; 3) implementation instructions to select and tailor recommendations; 4) alternative formatting of the recommendations; 5) data collection worksheets; and 6) research and methodological evidence supporting the toolkit’s content.

**Table 3** summarizes the quantitative survey ratings related to the toolkit’s recommendation statements, content, organization, and ease of implementation. Overall, all four areas of target stakeholder group recommendations received favorable mean ratings (range: 1.6-1.8). The mean ratings across domains ranged from 1.5 to 2.4, with the most and least favorable ratings attributed to the organization of recommendations and ease of implementation, respectively. The qualitative open-ended comments of dLVAD-AT members support the importance of simplifying recommendation wording to maximize toolkit uptake and the need for additional resources to assist in prioritizing and delineating concrete processes for implementing the recommendations.

**Table 3.**
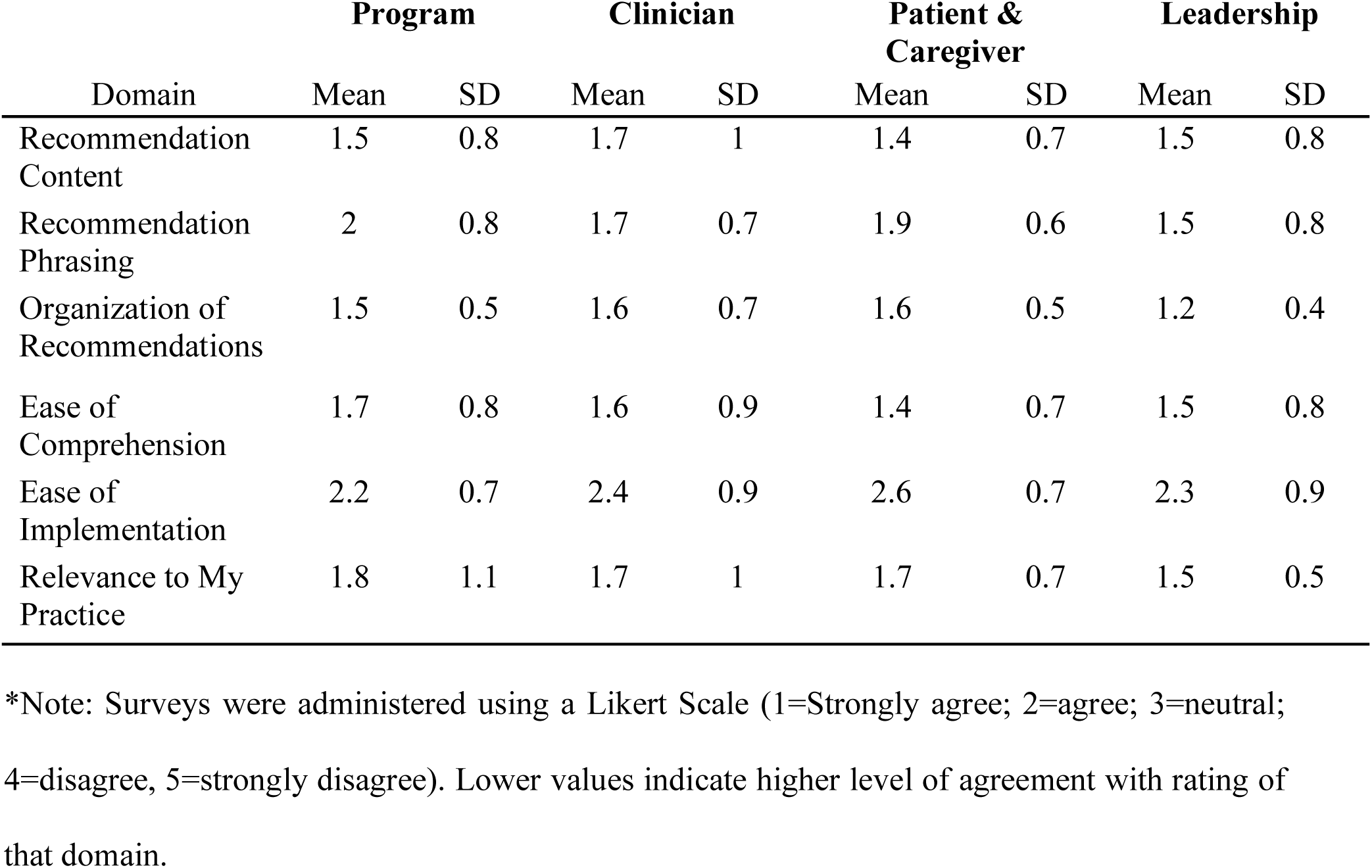
Summary Ratings of Toolkit Recommendations.

|  | **Program** |  | **Clinician** |  | **Patient & Caregiver** |  | **Leadership** |  |
| --- | --- | --- | --- | --- | --- | --- | --- | --- |
| Domain | Mean | SD | Mean | SD | Mean | SD | Mean | SD |
| Recommendation Content | 1.5 | 0.8 | 1.7 | 1 | 1.4 | 0.7 | 1.5 | 0.8 |
| Recommendation Phrasing | 2 | 0.8 | 1.7 | 0.7 | 1.9 | 0.6 | 1.5 | 0.8 |
| Organization of Recommendations | 1.5 | 0.5 | 1.6 | 0.7 | 1.6 | 0.5 | 1.2 | 0.4 |
| Ease of Comprehension | 1.7 | 0.8 | 1.6 | 0.9 | 1.4 | 0.7 | 1.5 | 0.8 |
| Ease of Implementation | 2.2 | 0.7 | 2.4 | 0.9 | 2.6 | 0.7 | 2.3 | 0.9 |
| Relevance to My Practice | 1.8 | 1.1 | 1.7 | 1 | 1.7 | 0.7 | 1.5 | 0.5 |

\*Note: Surveys were administered using a Likert Scale (1=Strongly agree; 2=agree; 3=neutral;
4=disagree, 5=strongly disagree). Lower values indicate higher level of agreement with rating of
that domain.

The final version of the toolkit contains 39 infection prevention recommendations. The recommendations span topics for four audiences: the VAD program (e.g., real-time provider communication), clinicians (e.g., multidisciplinary protocol development), patients & caregivers (e.g., engaging patient advisors in patient education), and VAD leadership (e.g., unit and service level data reporting). The packaging of the recommendations for the four audiences is intended to support programmatic priorities, resources, and implementation strategies to maximize the effectiveness of selected recommendations.

Based on the stakeholder feedback, the toolkit includes a customizable, eight-step implementation guide (**Figure 2)**. Steps 1-3 focus on assessing a VAD program’s strengths and weaknesses concerning infection prevention, identifying candidate recommendations that address areas of weakness, and providing tailorable criteria to prioritize which recommendations to implement. Steps 4-6 focus on identifying the roles and responsibilities of VAD team members during the implementation of the recommendations, developing a plan of action, and collecting discrete data elements to evaluate the implementation intervention. Steps 7-8 focus on implementing the action plan with periodic monitoring and reporting back to the VAD team. The toolkit has been designed to support the implementation of selected recommendations over iterative intervention cycles.

**Figure 2.**
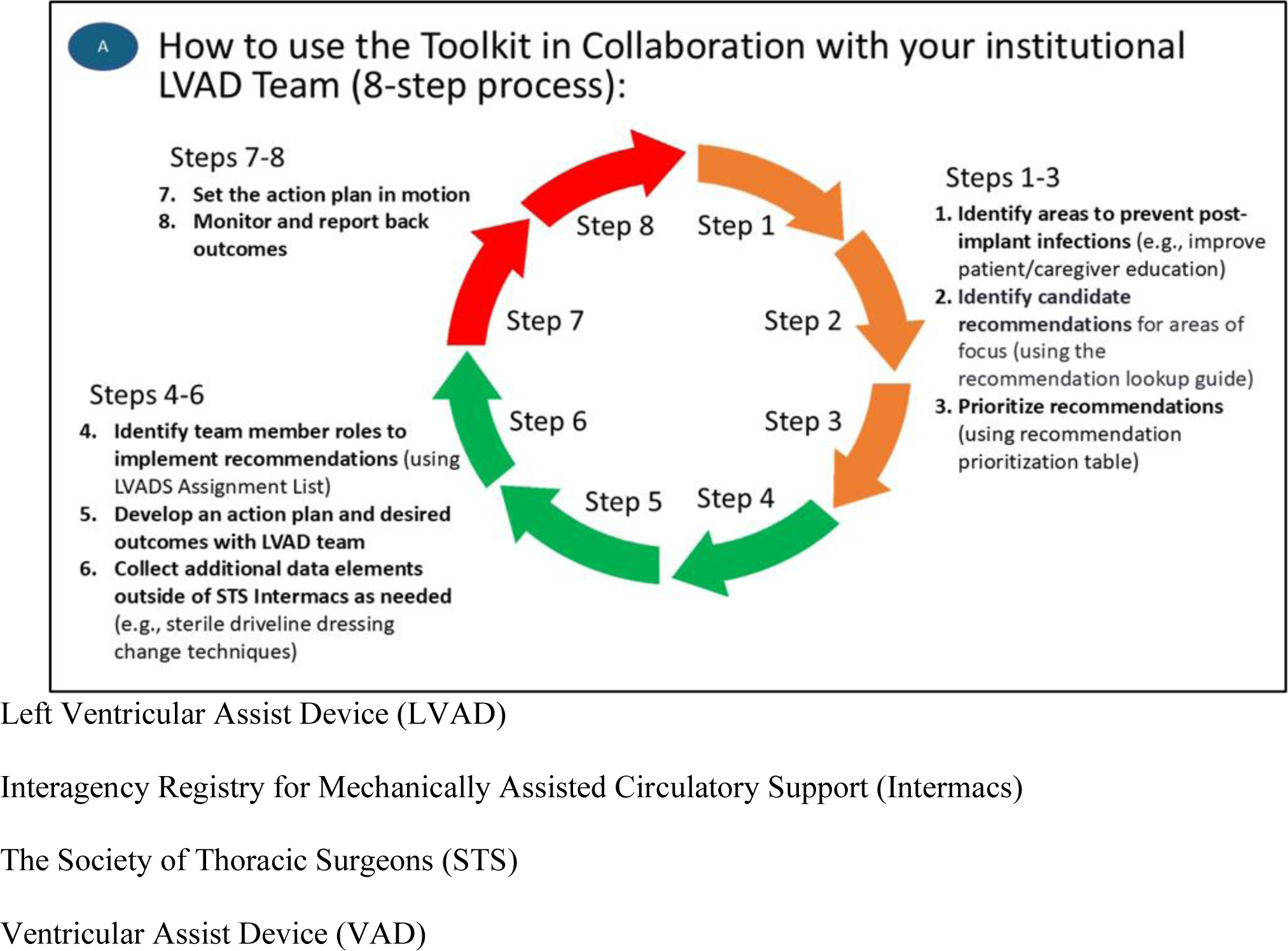
Eight-step process of the toolkit.

The toolkit includes accompanying resources to support the implementation of the recommendations. The resources (e.g., fact sheets regarding the rate and impact of infections, interactive worksheets) have been designed to support team discussions and the planning, implementing, and evaluating of hospital-specific action plans. For example, the toolkit resources include a prioritization table to support team-based decision-making regarding choosing recommendations. This guide enables teams to rate the anticipated impact and ease of implementation of each recommendation - a key gap noted during dLVAD-AT focus groups. Other implementation materials include a L-V-A-D-S assignment matrix that supports the delegation of roles and tasks to VAD team members: <u>L</u>eader (e.g., administrative and institutional support), <u>V</u>alidator (e.g., data navigation training), <u>A</u>ctor (e.g., educational lead), <u>D</u>elegator (task delegation authority), <u>S</u>upporter (e.g., data gathering support). These resources have been developed to support the implementation, evaluation, and anticipated effectiveness of hospital-specific infection prevention initiatives.

### Pilot Feedback on the Toolkit

Two VAD team members practicing at a high-volume dLVAD implanting institution piloted the toolkit. The study team received qualitative feedback, which indicated preliminary evidence of the toolkit’s acceptability regarding the feasibility of implementing the toolkit within their VAD programs. Overall, VAD team members noted the importance of the toolkit’s collaborative engagement materials and customizable implementation support. One respondent shared, “It’s helpful to have a step-by-step guide to help programs work through the infection prevention/management process. It can be hard to know where to start.” Respondents favorably commented on the ability to tailor the implementation of the recommendations: “I appreciate how you mention this should be tailored to each individual program based on context. I think it’s important to highlight it’s not one-size-fits-all.” However, respondents shared concerns about the toolkit’s length and complex organizational structure that may inhibit its use, especially for busy clinicians. Accordingly, one respondent offered suggestions to condense the toolkit to make it “easy to navigate, concise, and use. r friendly.” The study team received additional suggestions to refine the wording of the recommendation statements. The analytical team reviewed the feedback and responsively advanced the toolkit’s anticipated effectiveness by creating an interactive website to disseminate the toolkit, **eFigure 2**.

## DISCUSSION

This large-scale, multistage, mixed methods study advances determinants of and targeted action plans for reducing infections following dLVAD implantation. This study makes the following three important contributions. First, this study is among the first to leverage a multistage mixed methods research design to evaluate determinants of infections in this setting. The integration of qualitative and quantitative findings supports the development of contextualized and practice-informed infection prevention recommendations. Second, this study is among the first to develop evidence-informed infection prevention recommendations that span the acute and longer-term post-implantation periods. Third, this study is among the first to leverage focus groups with VAD team stakeholders to design implementation materials to advance local adoption.

More than 40%^1^ of patients undergoing dLVAD are at risk for developing a spectrum of infections within 1 year of implantation, reaching nearly 60%^2^ within 2 years. Patients developing infections in this setting are, in turn, exposed to an increased risk for further morbidity and mortality, as well as decrements in quality of life.^4,6^ Large-scale interhospital variability in post-implantation infections is unaccounted for by differences in patient case mix.^3^ Fragmentation within a hospital’s care delivery network (defined through network analyses to include the number of shared patients among clinicians) has been identified as a significant predictor of outcomes, including infections and mortality.^18^

To date, many infection prevention initiatives have been limited in how they incorporate stakeholder input.^7,11,19^ The present study integrated qualitative and quantitative findings to design a customizable infection prevention toolkit. The formative mixed methods assessment stage leveraged quantitative analyses of merged clinical registry datasets to select candidate hospitals (based in part on their risk-adjusted 180-day infection rates) for qualitative inquiry. Next, infection prevention themes emerging from the qualitative analysis of stakeholder interviews were merged with quantitative findings to identify emerging themes that served as the foundation for an infection prevention toolkit. Last, the qualitative and quantitative data generated from surveys and focus groups with our dLVAD-AT members supported the development of iteratively enhanced recommendations and implementation resources.

Prior structured literature reviews and surveys across dLVAD practices have identified risk factors for and interventions to prevent infections. Broadly, these studies have focused on: (1) patient risk factors^9,20^ (e.g., obesity, malnutrition, hypoalbuminemia); (2) non-patient factors^8^ (e.g., increasing center experience); (3) intraoperative strategies^8,21^ (e.g., silicone-skin interface at the driveline exit site); (4) diagnosis and early treatment of infections^20^ (e.g., use of F-fluorodeoxyglucose positron emission tomography, serial surgical debridement); and (5) driveline care^22^ (e.g., driveline immobilization). While findings from these studies inform which practices “work”, they lack insight into barriers and facilitators for advancing their implementation (e.g., patients may lack a stable caregiver to facilitate sterile driveline dressing changes, patients may have inadequate access to sterile dressing change kits). The present study’s qualitative interviews with key stakeholders across low and high-performing hospitals revealed the role of a VAD team’s structure and function to (1) address barriers for advancing evidence-based practices and (2) support quality assurance, standardization of practices, and targeted improvement. Uniquely, the present study advances the development of a toolkit, with content and recommendations specific to the VAD program (e.g., real-time provider communication), clinicians (e.g., multidisciplinary protocol development), patients & caregivers (e.g., engaging patient advisors in patient education), and VAD leadership (e.g., unit and service level data reporting). Notably, the resources contained within the toolkit (e.g., recommendation prioritization table) enable VAD program team members to prioritize recommendations based on each hospital’s specific context (e.g., resources and staffing) and required resources for implementing the recommendations.

Solely publishing findings that support evidence-based infection practices is insufficient for advancing their everyday use across diverse healthcare contexts. The field of implementation science^23^ has defined the strategies (e.g., promoting adaptability and engaging key stakeholders) necessary to advance the incorporation of evidence-informed practices within healthcare delivery systems. The present study leveraged a stakeholder advisory committee composed of VAD-team members (e.g., cardiac surgeons, cardiologists, VAD Coordinators, infectious disease personnel) to identify key facets of the toolkit’s content, resources, and dissemination plan to facilitate its adaptability and use across diverse settings. This feedback was leveraged to iteratively enhance the toolkit’s content and resources (e.g., an 8-step implementation guide) to promote its implementation across VAD programs. The prototype toolkit was further piloted and disseminated online to enhance its usability and anticipated effectiveness.

This study’s findings should be considered in light of several limitations. First, while the toolkit was developed by key clinical and non-clinical VAD stakeholders, further diversity of perspectives may enhance the toolkit’s anticipated effectiveness. Second, while this large mixed methods study leveraged a national advisory group and pilot testing to advance the toolkit’s usability and acceptability, further evidence (e.g., acceptability, appropriateness, feasibility) across VAD programs is warranted.

This multistage mixed methods study integrated national registry and claims-based data on dLVADs with qualitative interviews to develop an adaptable infection prevention toolkit. The toolkit includes accompanying resources as well as implementation and dissemination materials to support its use. Future work should evaluate the effectiveness of implementing this infection prevention toolkit across diverse dLVAD settings. More broadly, the integration of stakeholder feedback into the development and design of a customizable toolkit may be used as a methodological approach for other high-risk operations.

## Data Availability

The study authors will adhere to existing regulatory requirements. Raw data files are restricted by data use agreements, but summary statistics can be requested and will be shared after the study team reviews and approves the request.

## ACKNOWLEDGMENTS

We wish to extend our heartfelt gratitude to our esteemed colleague, mentor, and friend, Dr. Michael D. Fetters, for his invaluable contributions as a past collaborator. Dr. Fetters’ boundless insights and unwavering passion for discovery and innovation have profoundly shaped our research, leaving an indelible mark on our personal and professional journeys. The authors acknowledge the support from the Michigan Society of Thoracic and Cardiovascular Surgeons Quality Collaborative (MSTCVS-QC), as well as Blue Cross and Blue Shield of Michigan and Blue Care Network (BCBSM), which are part of the BCBSM Value Partnerships program. The content is solely the authors’ responsibility and does not represent the official views of the MSTCVS-QC, Blue Cross Blue Shield of Michigan and Blue Care Network, or any of its employees.

## Online Supplemental Material

**Table S1.**
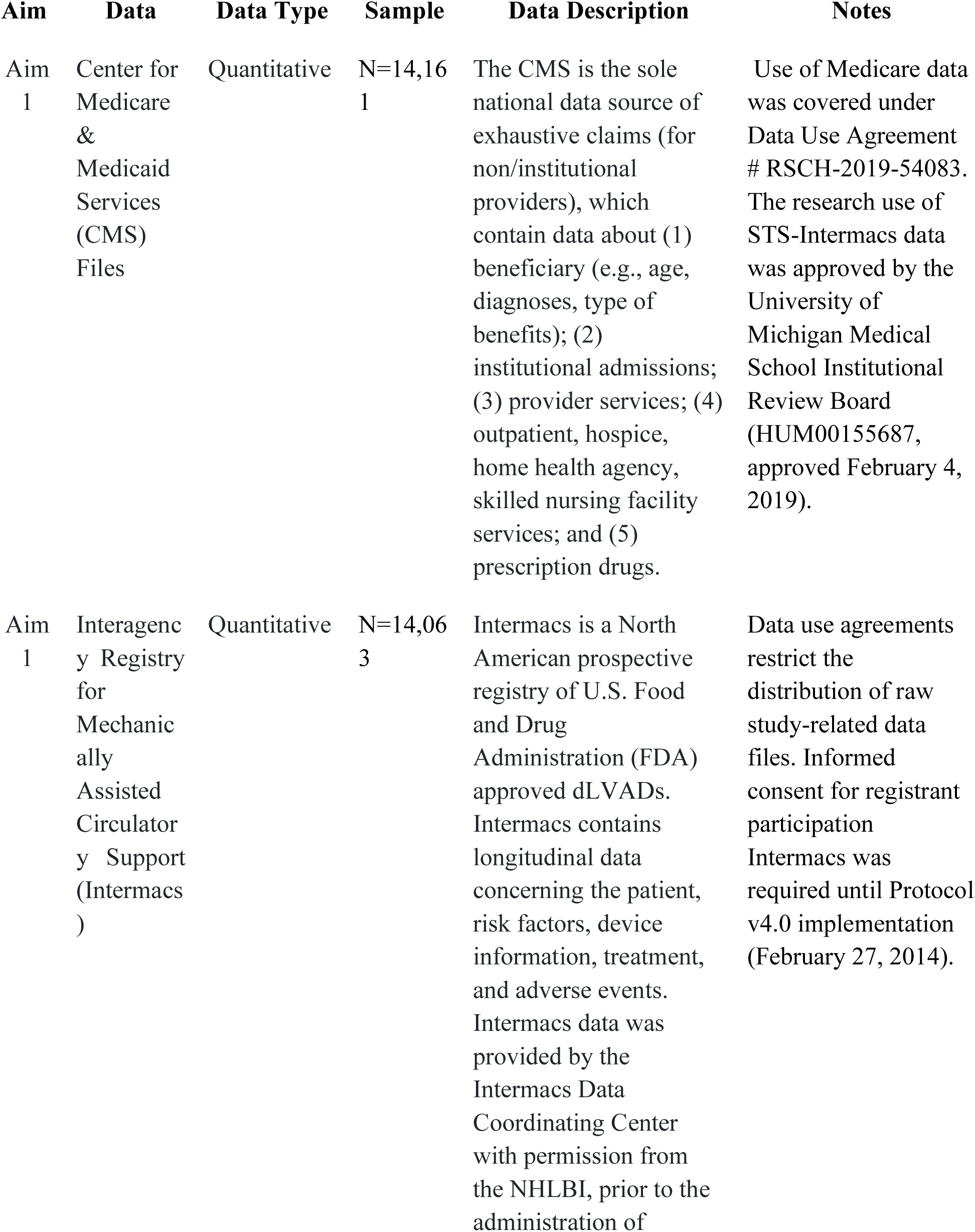
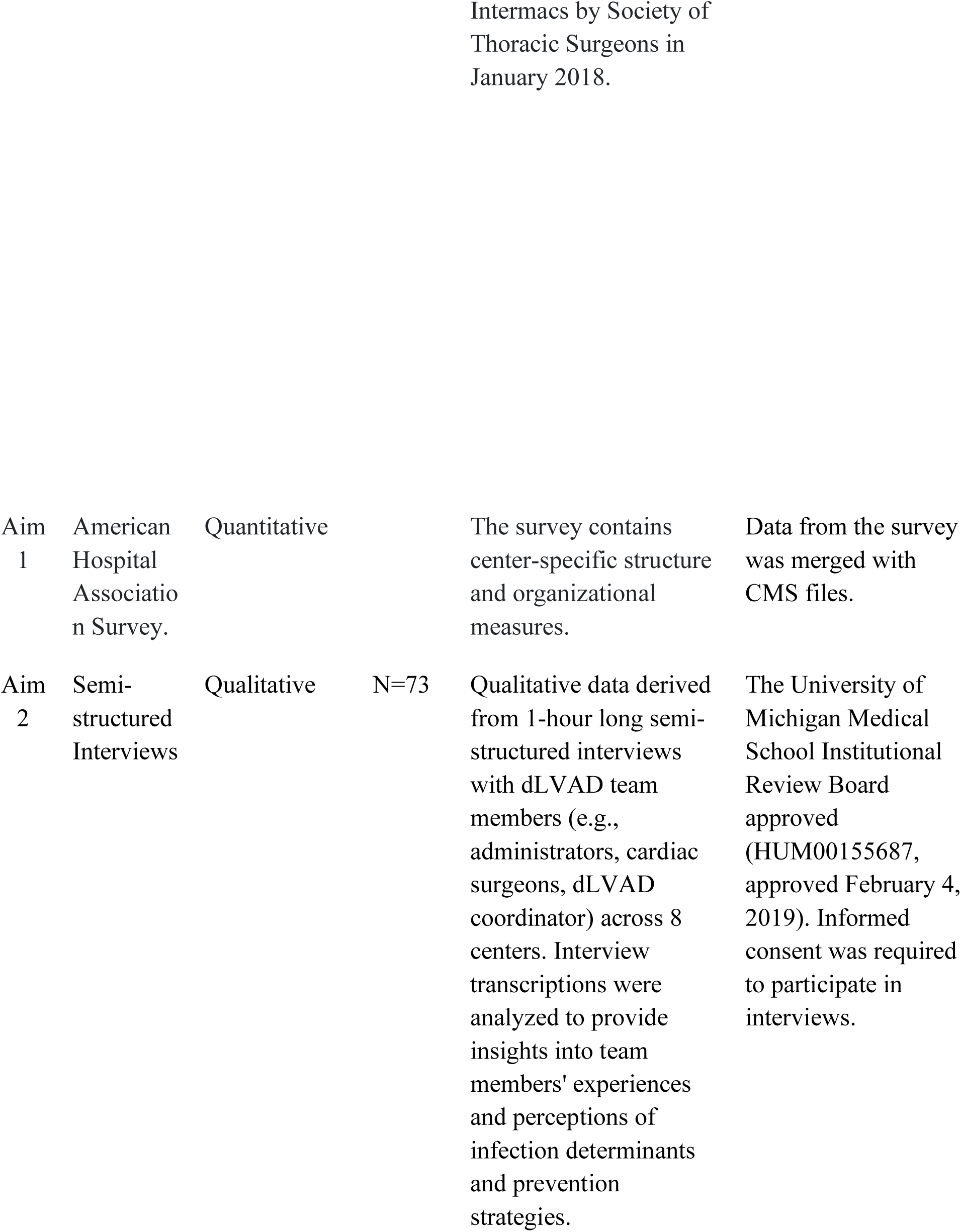
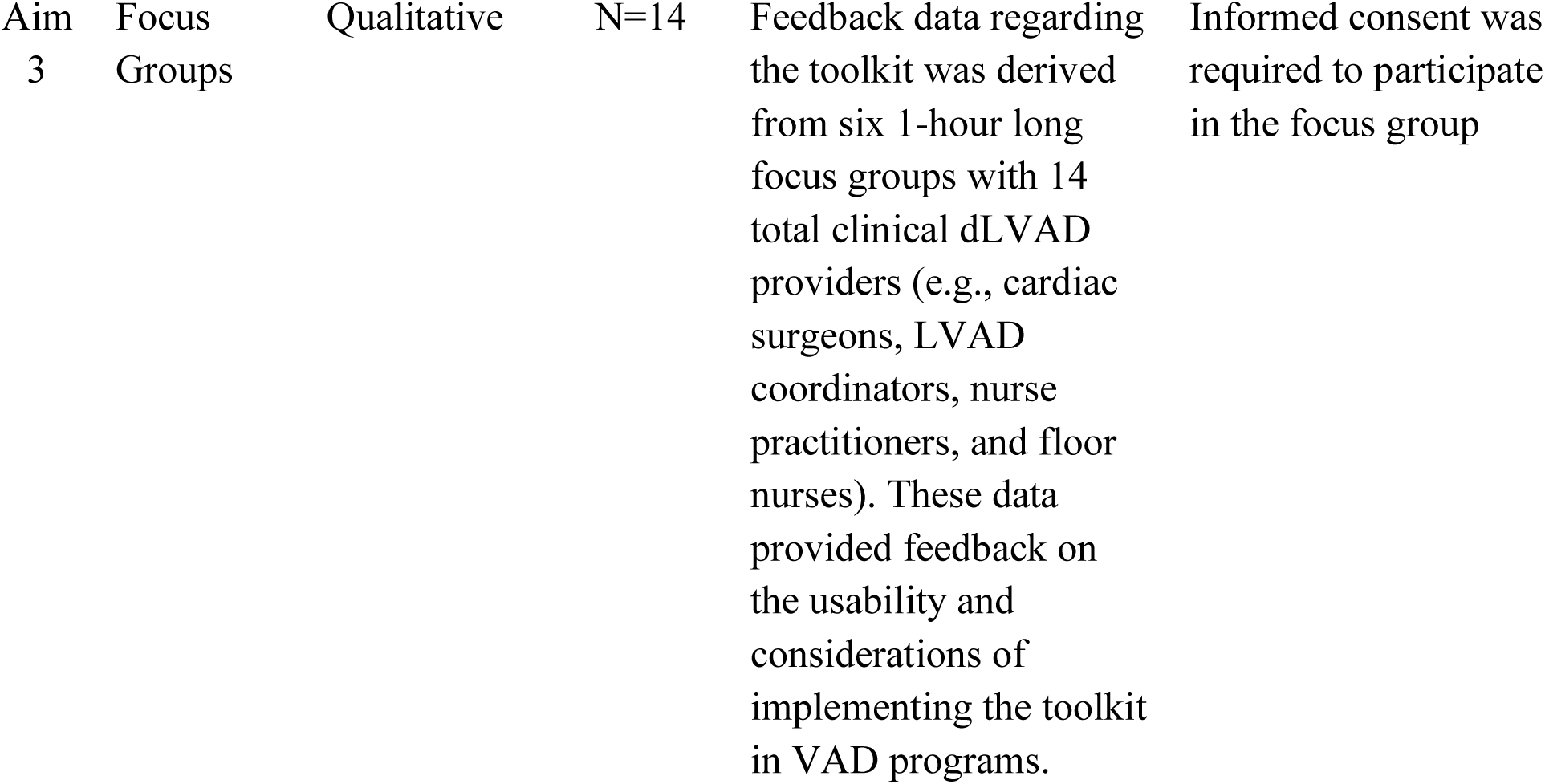
Data Sources.

| **Aim** | **Data** | **Data Type** | **Sample** | **Data Description** | **Notes** |
| --- | --- | --- | --- | --- | --- |
| Aim 1 | Center for Medicare & Medicaid Services (CMS) Files | Quantitative | N=14,161 | The CMS is the sole national data source of exhaustive claims (for non/institutional providers), which contain data about (1) beneficiary (e.g., age, diagnoses, type of benefits); (2) institutional admissions; (3) provider services; (4) outpatient, hospice, home health agency, skilled nursing facility services; and (5) prescription drugs. | Use of Medicare data was covered under Data Use Agreement # RSCH-2019-54083. The research use of STS-Intermacs data was approved by the University of Michigan Medical School Institutional Review Board (HUM00155687, approved February 4, 2019). |
| Aim 1 | Interagency Registry for Mechanically Assisted Circulatory Support (Intermacs) | Quantitative | N=14,063 | Intermacs is a North American prospective registry of U.S. Food and Drug Administration (FDA) approved dLVADs. Intermacs contains longitudinal data concerning the patient, risk factors, device information, treatment, and adverse events. Intermacs data was provided by the Intermacs Data Coordinating Center with permission from the NHLBI, prior to the administration of | Data use agreements restrict the distribution of raw study-related data files. Informed consent for registrant participation Intermacs was required until Protocol v4.0 implementation (February 27, 2014). |
| Aim<br>1 | American<br>Hospital<br>Associatio<br>n Survey. | Quantitative |  | The survey contains<br>center-specific structure<br>and organizational<br>measures. | Data from the survey<br>was merged with<br>CMS files. |
| Aim<br>2 | Semi-structured<br>Interviews | Qualitative | N=73 | Qualitative data derived<br>from 1-hour long semi-structured interviews<br>with dLVAD team<br>members (e.g.,<br>administrators, cardiac<br>surgeons, dLVAD<br>coordinator) across 8<br>centers. Interview<br>transcriptions were<br>analyzed to provide<br>insights into team<br>members' experiences<br>and perceptions of<br>infection determinants<br>and prevention<br>strategies. | The University of<br>Michigan Medical<br>School Institutional<br>Review Board<br>approved<br>(HUM00155687,<br>approved February 4,<br>2019). Informed<br>consent was required<br>to participate in<br>interviews. |
| Aim<br>3 | Focus<br>Groups | Qualitative | N=14 | Feedback data regarding the toolkit was derived from six 1-hour long focus groups with 14 total clinical dLVAD providers (e.g., cardiac surgeons, LVAD coordinators, nurse practitioners, and floor nurses). These data provided feedback on the usability and considerations of implementing the toolkit in VAD programs. | Informed consent was required to participate in the focus group |

**Table S2.**
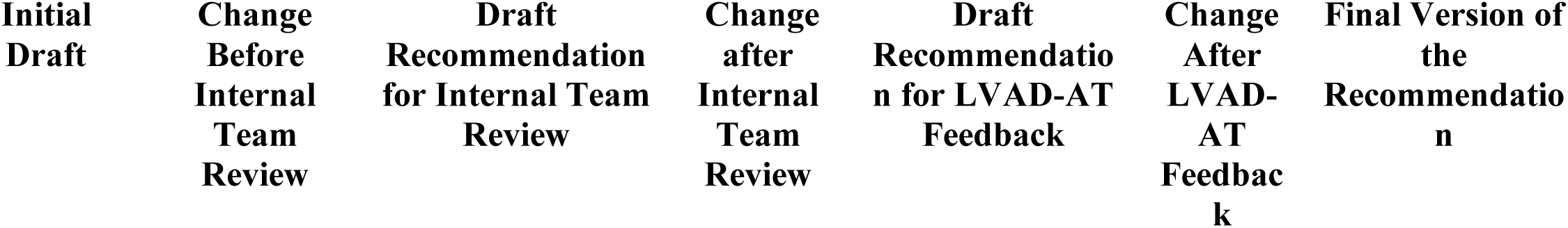
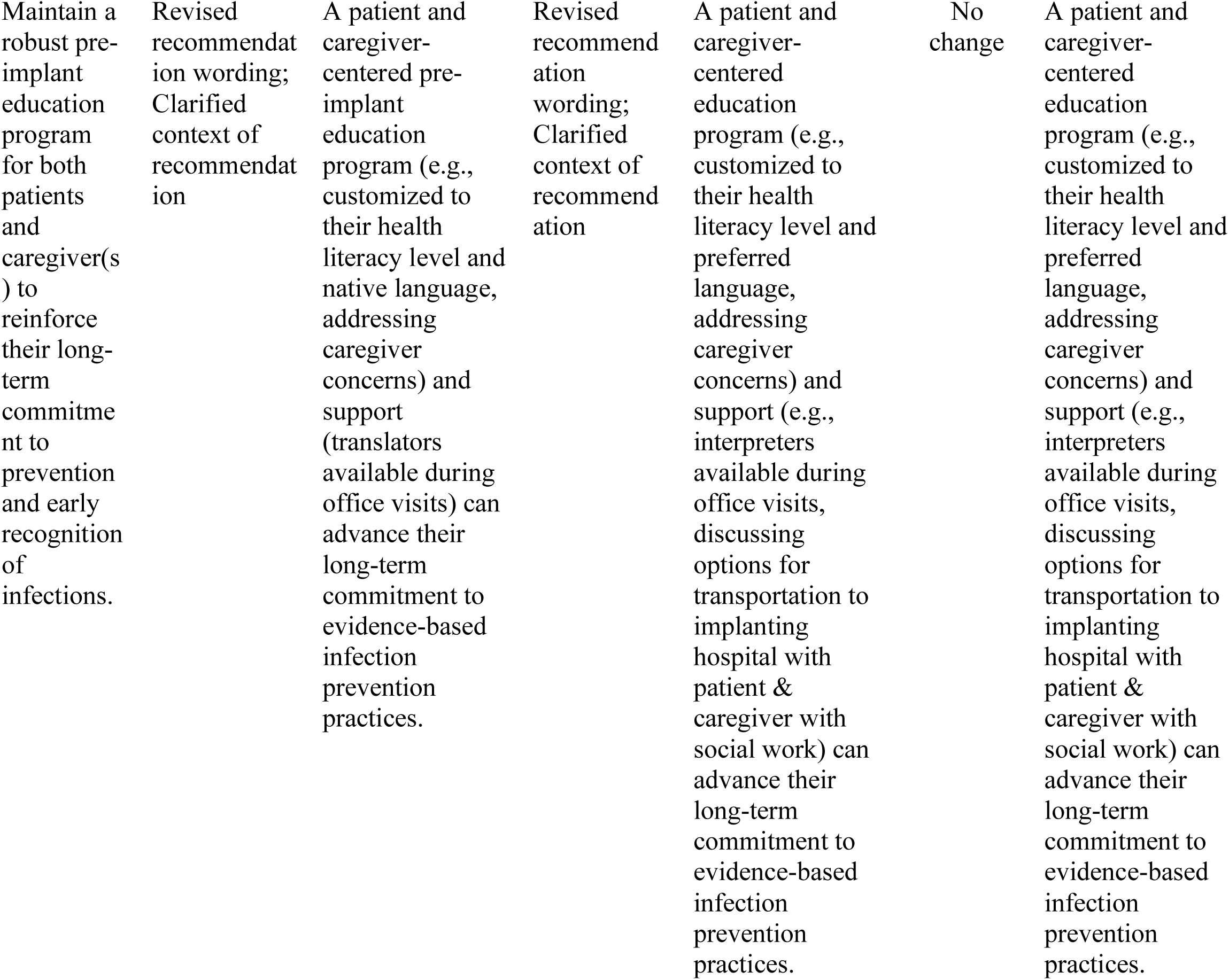
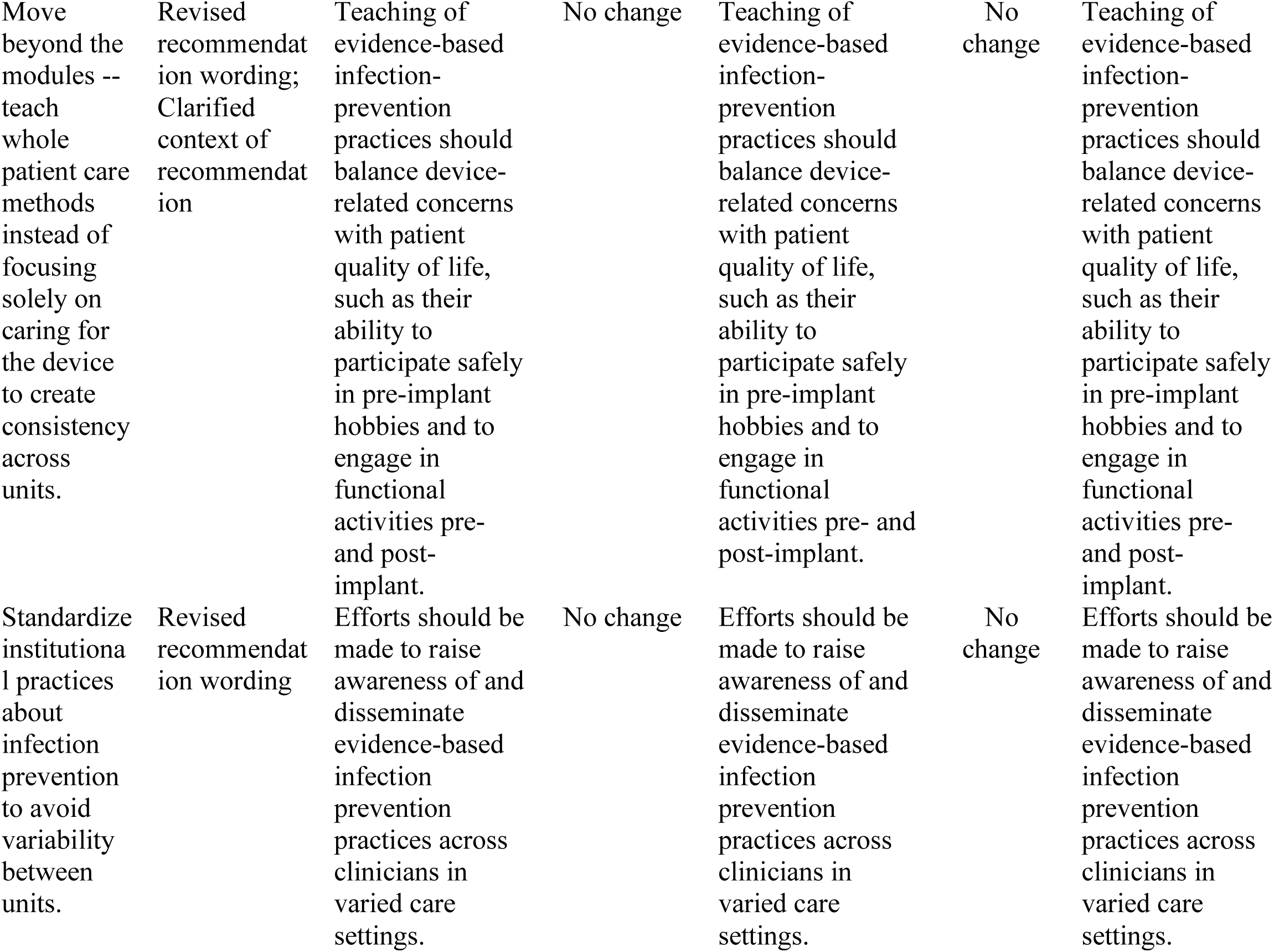
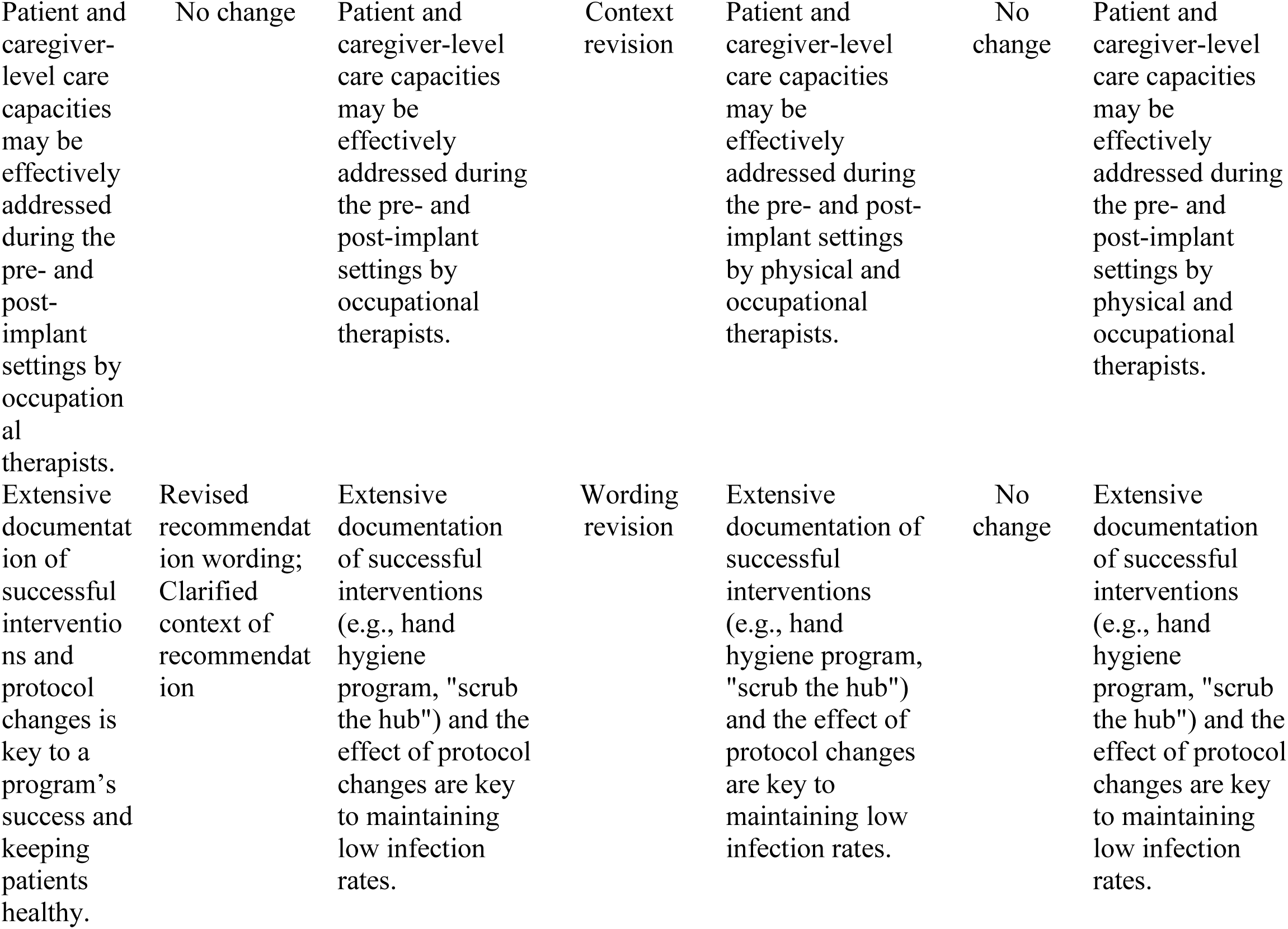
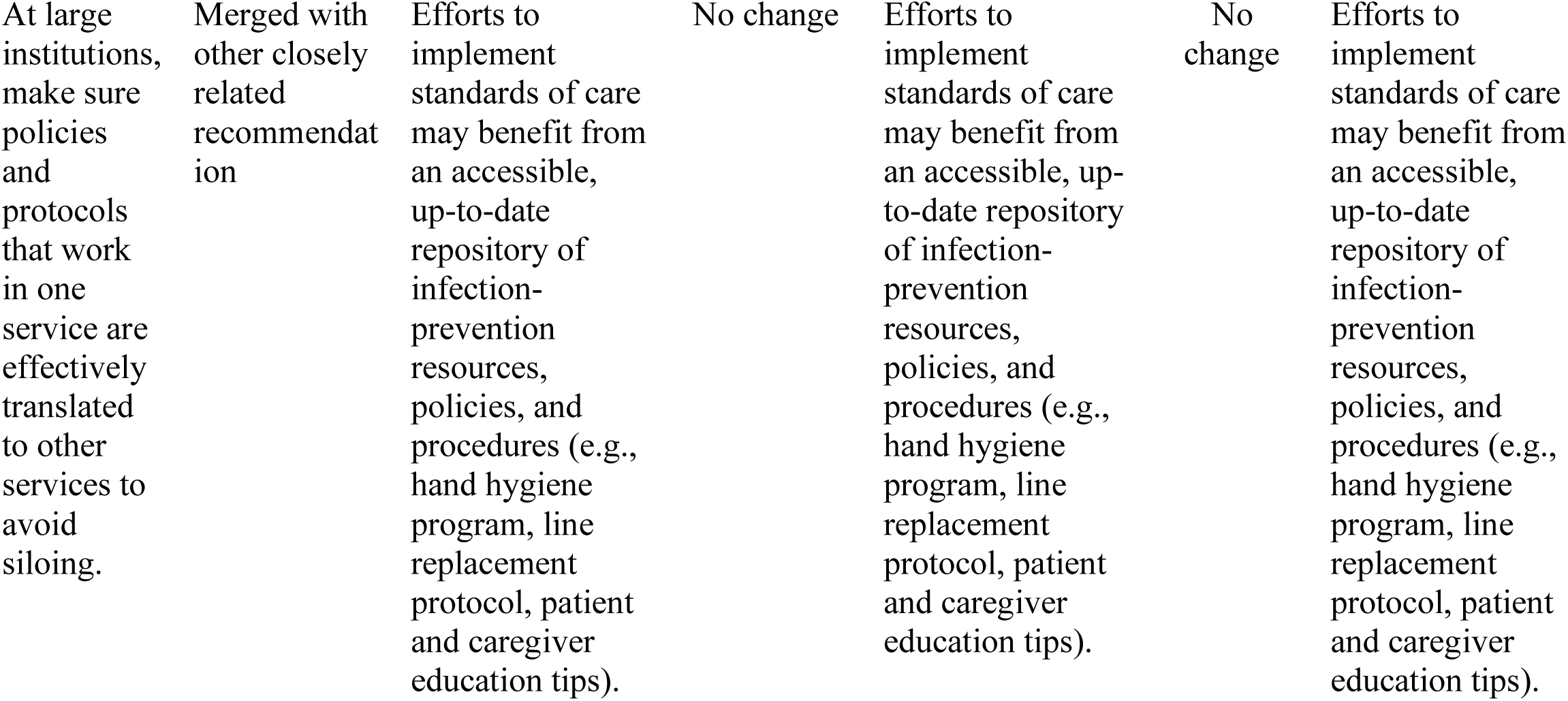
Revisions Made to Toolkit Recommendations.

**Figure S1.**
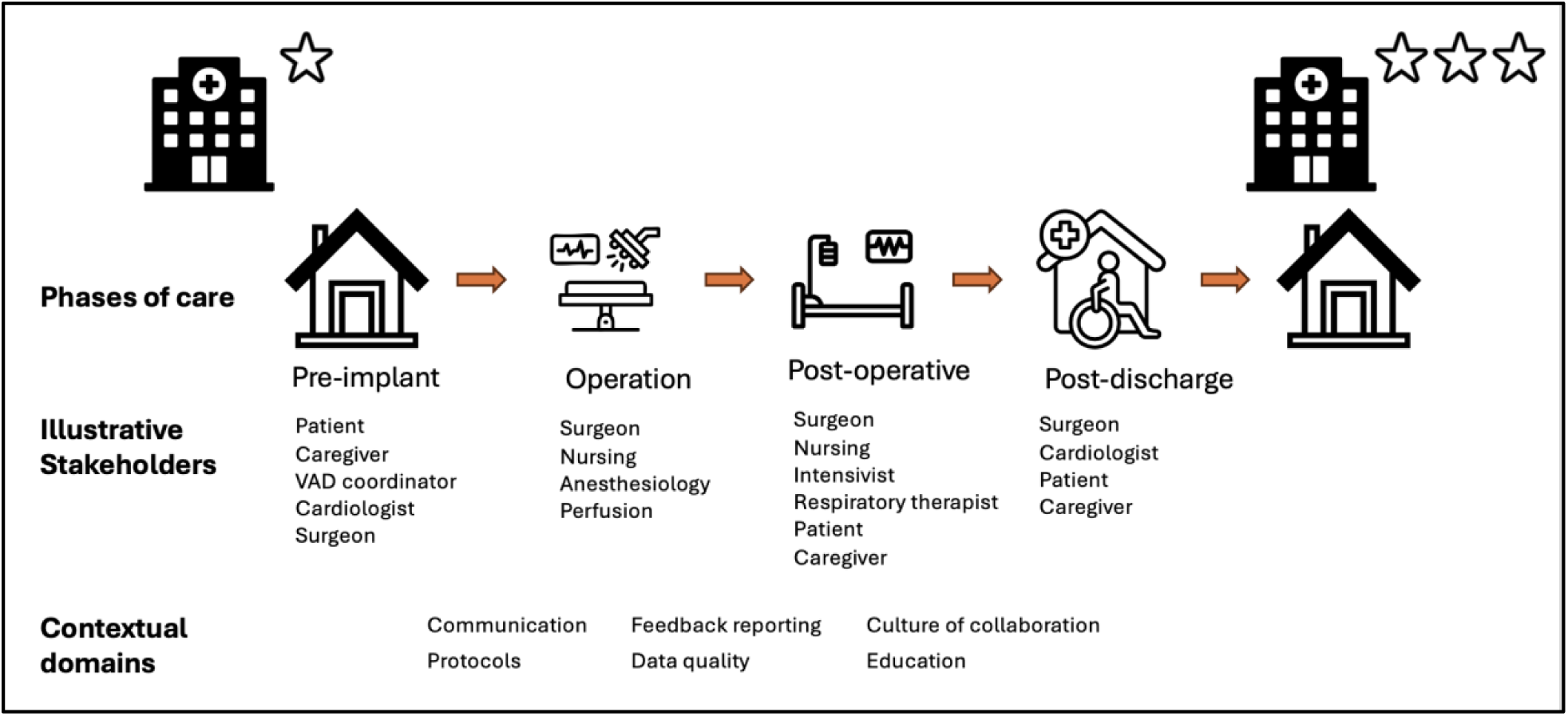
Conceptual Framework of dLVAD care.

**Figure S2.**
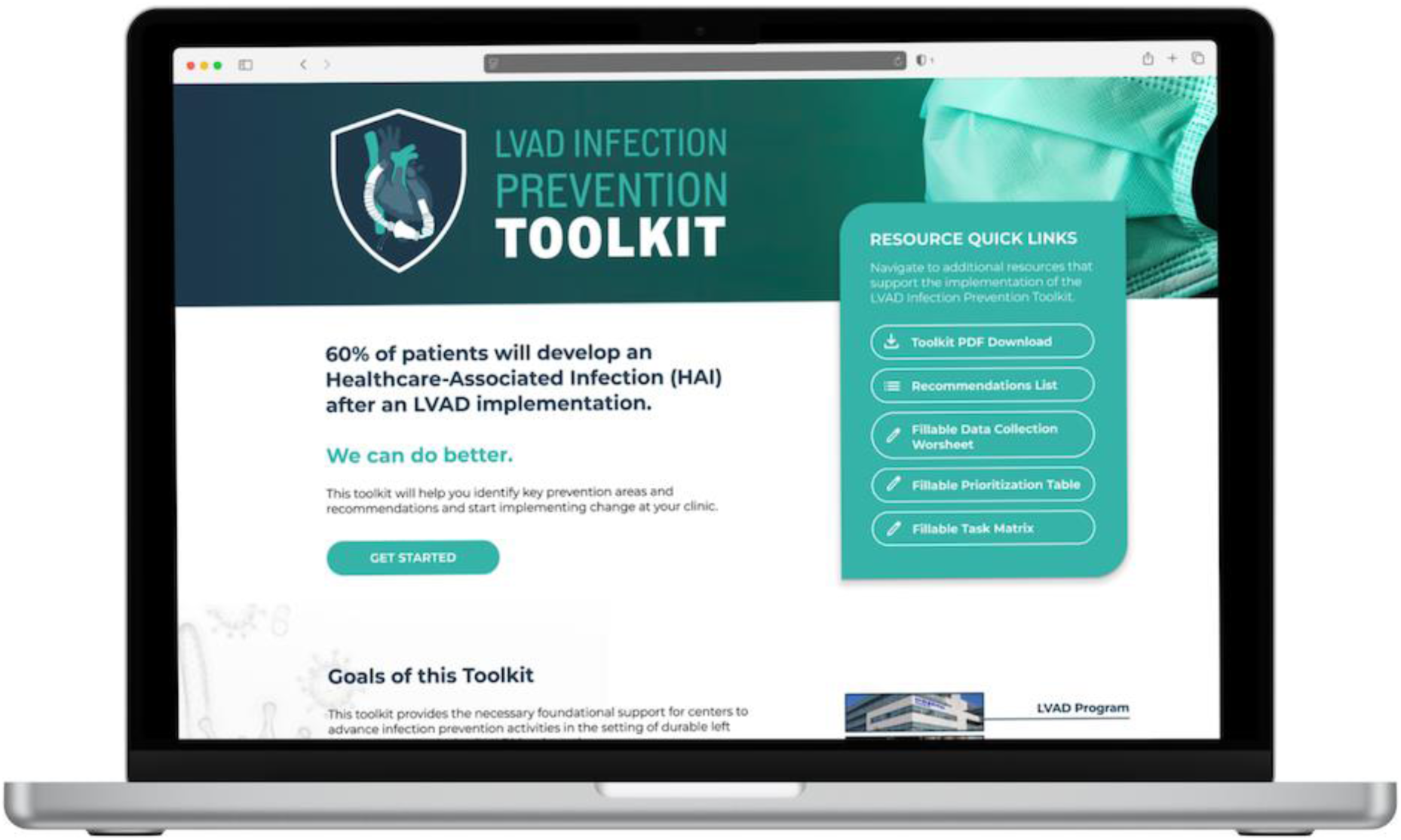
Selected Screenshot of the Infection Prevention Toolkit.

## Non-Standard Abbreviations and Acronyms

dLVAD: Durable Left Ventricular Assist Device
Intermacs: Interagency Registry for Mechanically Assisted Circulatory Support
STS: The Society of Thoracic Surgeons
VAD: Ventricular Assist Device

## REFERENCES

1. Pienta M, Shore S, Pagani FD, Likosky DS, Likosky DS, Pagani FD, Shaaban A, Abou El Ela AA, Tang PC, Thompson MP, Aaronson K, Shore S, Cascino T, Salciccioli KB, Zhang M, McCullough JS, Hou M, Janda AM, Mathis MR, Watt TMF, Pienta MJ, Brescia A, Airhart A, Liesman D, Nassar K. Rates and types of infections in left ventricular assist device recipients: A scoping review. JTCVS Open. 2021;8:405–411.

2. Patel CB, Blue L, Cagliostro B, Bailey SH, Entwistle JW, John R, Thohan V, Cleveland JC Jr, Goldstein DJ, Uriel N, Su X, Somo SI, Sood P, Mehra MR. Left ventricular assist systems and infection-related outcomes: A comprehensive analysis of the MOMENTUM 3 trial. J Heart Lung Transplant [Internet]. 2020;Available from: 10.1016/j.healun.2020.03.002

3. Likosky DS, Yang G, Zhang M, Malani PN, Fetters MD, Strobel RJ, Chenoweth CE, Hou H, Pagani FD, Michigan Congestive Heart Failure Investigators. Interhospital variability in health care-associated infections and payments after durable ventricular assist device implant among Medicare beneficiaries. J Thorac Cardiovasc Surg. 2022;164:1561–1568.

4. Zhou S, Yang G, Zhang M, Pienta M, Chenoweth CE, Pagani FD, Aaronson KD, Fetters MD, Chandanabhumma PP, Cabrera L, Hou H, Malani PN, Likosky DS, Michigan Congestive Heart Failure Investigators. Mortality following durable left ventricular assist device implantation by timing and type of first infection. J Thorac Cardiovasc Surg [Internet]. 2021;Available from: 10.1016/j.jtcvs.2021.10.056

5. Yang G, Zhang M, Zhou S, Hou H, Grady KL, Stewart JW 2nd, Chenoweth CE, Aaronson KD, Fetters MD, Chandanabhumma PP, Pienta MJ, Malani PN, Hider AM, Cabrera L, Pagani FD, Likosky DS, Michigan Congestive Heart Failure Investigators. Incompleteness of health-related quality of life assessments before left ventricular assist device implant: A novel quality metric. J Heart Lung Transplant. 2022;41:1520–1528.

6. Zhou S, Yang G, Hou H, Zhang M, Grady KL, Chenoweth CE, Aaronson KD, Pienta M, Fetters MD, Paul Chandanabhumma P, Stewart JW 2nd, Cabrera L, Malani PN, Pagani FD, Likosky DS, Michigan Congestive Heart Failure Investigators. Infections following left ventricular assist device implantation and 1-year health-related quality of life. J Heart Lung Transplant. 2023;42:1307–1315.

7. Chandanabhumma PP, Zhou S, Fetters MD, Likosky DS. Expanding Our Methodological Toolbox to Improve Quality: The Role of Mixed-Methods Evaluations. Circ Cardiovasc Qual Outcomes. 2023;16:e009629.

8. Shore S, Pienta MJ, Watt TMF, Yost G, Townsend WA, Cabrera L, Fetters MD, Chenoweth C, Aaronson KD, Pagani FD, Likosky DS. Non-patient factors associated with infections in LVAD recipients: A scoping review. J Heart Lung Transplant. 2022;41:1–16.

9. Pienta MJ, Shore S, Watt TMF, Yost G, Townsend W, Cabrera L, Fetters MD, Chenoweth C, Aaronson K, Pagani FD, Likosky DS, Michigan Congestive Heart Failure Investigators. Patient factors associated with left ventricular assist device infections: A scoping review. J Heart Lung Transplant. 2022;41:425–433.

10. Tracer methodology fact sheet [Internet]. [cited 2024 Jan 31];Available from: https://www.jointcommission.org/resources/news-and-multimedia/fact-sheets/facts-about-tracer-methodology/

11. Chandanabhumma PP, Swaminathan S, Cabrera LM, Hou H, Yang G, Kim KD, Janda AM, Nassar K, Malani PN, Zhang M, Funk RJ, Aaronson KD, Wu J, Pagani FD, Likosky DS. Enhancing qualitative and quantitative data linkages in complex mixed methods designs: Illustrations from a multi-phase healthcare delivery study. J Mix Methods Res. 2024;18:235–246.

12. Chandanabhumma PP, Fetters MD, Pagani FD, Malani PN, Hollingsworth JM, Funk RJ, Aaronson KD, Zhang M, Kormos RL, Chenoweth CE, Shore S, Watt TMF, Cabrera L, Likosky DS. Understanding and Addressing Variation in Health Care-Associated Infections After Durable Ventricular Assist Device Therapy: Protocol for a Mixed Methods Study. JMIR Res Protoc. 2020;9:e14701.

13. Fetters MD. The Mixed Methods Research Workbook: Activities for Designing, Implementing, and Publishing Projects. SAGE Publications; 2019.

14. Liang Q, Ward S, Pagani FD, Sinha SS, Zhang M, Kormos R, Aaronson KD, Althouse AD, Kirklin JK, Naftel D, Likosky DS. Linkage of Medicare Records to the Interagency Registry of Mechanically Assisted Circulatory Support. Ann Thorac Surg. 2018;105:1397– 1402.

15. King N, Brooks JM. Template analysis for business and management students. 1 Oliver’s Yard, 55 City Road London EC1Y 1SP: SAGE Publications Ltd; 2017.

16. Boyatzis RE. Transforming qualitative information. Thousand Oaks, CA: SAGE Publications; 1998.

17. Glaser BG, Strauss AL. The discovery of grounded theory. Routledge; 2017.

18. Funk RJ, Pagani FD, Hou H, Zhang M, Yang G, Malani PN, Chandanabhumma PP, Cabrera L, Kim KD, Likosky DS, Michigan Congestive Heart Failure Investigators. Care fragmentation predicts 90-day durable ventricular assist device outcomes. Am J Manag Care. 2022;28:e444–e451.

19. Hoefsmit PC, Schretlen S, Burchell G, van den Heuvel J, Bonjer J, Dahele M, Zandbergen R. Can Quality Improvement Methodologies Derived from Manufacturing Industry Improve Care in Cardiac Surgery? A Systematic Review. J Clin Med Res [Internet]. 2022;11. Available from: 10.3390/jcm11185350

20. Michelly Gonçalves Brandão S, Belletti Mutt Urasaki M, Machado Pires Lemos D, Neres Matos L, Takahashi M, Cristina Nogueira P, Lucia Conceição de Gouveia Santos V. Risk factors, diagnostic methods and treatment of infection in adult patients undergoing left ventricular assist device implantation: A scoping review. Intensive Crit Care Nurs. 2024;84:103726.

21. Brandão SMG, Urasaki MBM, Lemos DMP, Matos LN, Takahashi M, Nogueira PC, de Gouveia Santos VLC. Perioperative interventions for the prevention of surgical wound infection in adult patients undergoing left ventricular assist devices implantation: A scoping review. Intensive Crit Care Nurs. 2024;82:103658.

22. Krzelj K, Petricevic M, Gasparovic H, Biocina B, McGiffin D. Ventricular Assist Device Driveline Infections: A Systematic Review. Thorac Cardiovasc Surg. 2022;70:493–504.

23. Bauer MS, Damschroder L, Hagedorn H, Smith J, Kilbourne AM. An introduction to implementation science for the non-specialist. BMC Psychol. 2015;3:32.

